# COVID-19-related symptoms 6 months after the infection - Update on a prospective cohort study in Germany

**DOI:** 10.1101/2021.02.12.21251619

**Authors:** Barbara Rauch, Stefanie Kern-Matschilles, Stefanie J. Haschka, Vanessa Sacco, Anne L. Potzel, Friederike Banning, Irina Benz, Mandy Meisel, Jochen Seissler, Christina Gar, Andreas Lechner

## Abstract

**Objective:** Many anecdotal reports indicate the presence of ‘long COVID’ – COVID-19-related symptoms weeks to months after the acute illness. However, frequency and symptom-pattern of ‘long COVID’ in relation to acute disease severity are uncertain. As part of an ongoing, prospective cohort study we therefore conducted an online survey among adults 6 months after acute COVID-19.

**Methods:** The prospective online study Life&Covid is ongoing in Germany since May 2020. Participants were recruited 0 to 4 months after their SARS-CoV-2 infection und followed up by subsequent surveys. The survey 6 months after the infection was completed by 127 out of 148 individuals invited by email (86%). All grades of acute disease severity were included and 91% of the participants had been treated as outpatients during their acute illness.

**Results:** Six months after the infection, 67% of the study participants reported at least one symptom as a consequence of COVID-19. Exertional dyspnea (30% of participants), fatigue (25%) and diminished sense of taste/smell (19%) were the most common individual symptoms. At least one symptom, exertional dyspnea, and fatigue were reported more often after a severe acute illness, but diminished sense of taste/smell was unrelated to acute severity. Age group and sex did not associate with the frequency of symptoms at 6 months.

**Conclusions:** Based on this study, the prevalence of COVID-19-related symptoms 6 months after the infection is high. Some bias for overestimation may have affected this result. Nevertheless, ‘long COVID’ requires attention in medical care and a better scientific understanding.

## Introduction

Beyond the acute illness, a SARS-CoV-2 infection can cause prolonged symptoms, also termed ‘long COVID’ (1–3). However, the clinical course of ‘long COVID’ is heterogeneous (4) and may include diverse symptoms that range from mild to disabling. Furthermore, the underlying pathophysiology is unclear and probably complex (5).

From an epidemiologic standpoint, the frequency of ‘long COVID’ in relation to acute disease severity and the duration and nature of its predominant symptoms are still unclear. To address these questions, we are conducting a prospective cohort study after acute COVID-19 in Germany. Here, we report data from the 6-month survey of this study.

## Material and Methods

### Study design

This survey was part of the ongoing, prospective, online cohort study *Life&Covid* that is being conducted in Germany since May 2020. The study design has been described previously (6). In brief, the study mainly aimed at investigating the influence of lifestyle factors on the course of acute COVID-19. An additional aim was to record any long-term symptoms after the acute illness. The inclusion criteria for this study were a recent history of COVID-19 and an age of 18 years or older. Infection with SARS-CoV-2 had to be diagnosed by PCR from a nasopharyngeal swab or, in retrospect, by antibody testing. Additional inclusion criteria were permanent residence in Germany and online informed consent. Participants were recruited through medical information websites, social media postings and general news media. The study was approved by the ethics committee of the medical faculty of the Ludwig-Maximilians-Universität in Munich, Germany.

The time between the infection (February to June 2020) and study inclusion (May to July 2020) was between 0 and 4 months for each participant. We use the online tool Unipark (ww2.unipark.de) for this study to automate scheduled surveys at baseline, 3 months (omitted if overlapping with baseline survey), 6 and 12 months after the acute infection. At baseline, participants were able to decline to register for the follow-up surveys.

This manuscript reports data from the 127 participants of the 6-month survey. At baseline, 201 individuals were included in the study. Of these, 38 declined to register for the follow-up surveys and 4 had to be excluded from the follow-up because of duplicate email addresses. For another 11 participants, the email invitations for the 6-month survey were undeliverable and 21 did not complete the survey despite repeated invitations. In sum, 127 out of 148 invited individuals completed the 6-month survey (86%).

### Outcome variable

#### Participants were asked this multiple-choice question

“Do you still feel symptoms as a consequence of your COVID-19 illness? The link to COVID-19 does not have to be proven. It is sufficient if you believe a link exists.”

The possible answers were:

- Exertional dyspnoe
- Fatigue
- Diminished sense of taste/smell
- Difficulties in concentration
- Sleeplessness
- Headache
- Mood swings
- Vertigo/unsteady gait
- Cough
- Hyperhidrosis/fever/elevated body temperature
- Diarrhea
- Leg paresthesia/pain/numbness
- Rhinitis
- Painful breathing
- Other symptoms not listed here
- No symptoms

‘No symptoms’ was an exclusive answer. In all other cases, one or more answers were allowed. We report the proportion of individuals with at least on symptom and the proportions of individuals reporting each of the listed symptoms.

### Stratification variables

We also report stratified analyses for the proportion of individuals with at least one symptom and one of the 3 most common individual symptoms. For stratification, we applied sex, 3 age groups and 3 categories of acute COVID-19 severity.

### Statistical methods

Group comparisons were done with the Chi square test or the Fisher exact test, respectively. The sample size was determined by practical reasons. All eligible participants during the recruitment period were included. All questions of the online questionnaire had to be answered. Therefore, no data were missing.

## Results

The baseline data of the 127 study participants who completed the 6-month survey are summarized in **table 1**. For better estimates of the risk of overreporting of symptoms at 6 months, we compared these participants to the 38 participants who, at baseline, had declined to participate in the follow-up surveys. This comparison of baseline characteristics revealed no significant differences between the two groups. In particular, severity of the acute illness, proportion of outpatient treatment, reporting of at least one symptom as a consequence of COVID-19 in the baseline survey, age group and sex were distributed similarly (**table 1**).

**Table 1:** Baseline characteristics of the study participants

|  | Participants completing 6 months follow-up survey (n = 127) | Participants not consenting to follow-up (n = 38) | p-value |
| --- | --- | --- | --- |
| **Age group** |  |  |  |
| 18 – 39 years | 29 (22.8) | 14 (36.8) | 0.221 <sup>a</sup> |
| 40 – 59 years | 63 (49.6) | 16 (42.1) |  |
| ≥ 60 years | 35 (27.6) | 8 (21.1) |  |
| **Sex** |  |  |  |
| Female | 87 (68.5) | 26 (68.4) | 1.000 <sup>b</sup> |
| Male | 40 (31.5) | 12 (31.6) |  |
| **Severity of acute COVID-19** |  |  |  |
| Mild (asymptomatic or mild symptoms) | 39 (30.7) | 8 (21.1) | 0.511 <sup>a</sup> |
| Moderate (strong symptoms with fever or cough but no dyspnea) | 62 (48.8) | 21 (55.3) |  |
| Severe (strong symptoms including dyspnea, oxygen requirement, or mechanical ventilation) | 26 (20.5) | 9 (23.7) |  |
| **Method of SARS-CoV-2 testing** |  |  |  |
| PCR from nasopharyngeal swab | 102 (80.3) | 32 (84.2) | 0.813 <sup>b</sup> |
| Antibody testing (retrospective) | 25 (19.7) | 6 (15.8) |  |
| **Outpatient treatment during acute COVID-19** | 116 (91.3) | 33 (86.8) | 0.531 <sup>b</sup> |
| **Reporting at least one symptom as a consequence of COVID-19 in the baseline survey** | 74 (58.3) | 17 (44.7) | 0.193 <sup>b</sup> |
| **Preexisting, chronic pulmonary disease** | 12 (9.4) | 3 (7.9) | 1.000 <sup>b</sup> |
| **Preexisting, chronic cardiovascular disease** | 11 (8.7) | 0 (0.0) | 0.070 <sup>b</sup> |
| **Body mass index** |  |  |  |
| < 25 kg/m<sup>2</sup> | 73 (57.5) | 26 (68.4) | 0.261 <sup>b</sup> |
| ≥ 25 kg/m<sup>2</sup> | 54 (42.5) | 12 (31.6) |  |
| Data are n (%); group comparisons: <sup>a</sup> Chi square test, <sup>b</sup> Fisher exact test |  |  |  |

At 6 months, 67% of the study participants reported at least one symptom as a consequence of COVID-19. The 3 most common individual symptoms were exertional dyspnea (30%), fatigue (25%), and diminished sense of taste/smell (19%; **figure 1**). At least one reported symptom, exertional dyspnea, and fatigue were more common after severe acute illness, but this was not case for diminished sense of taste/smell (**figure 2**). Stratification by age group (**figure 3**) and sex (**figure 4**) revealed no significant differences.

**Figure 1:**
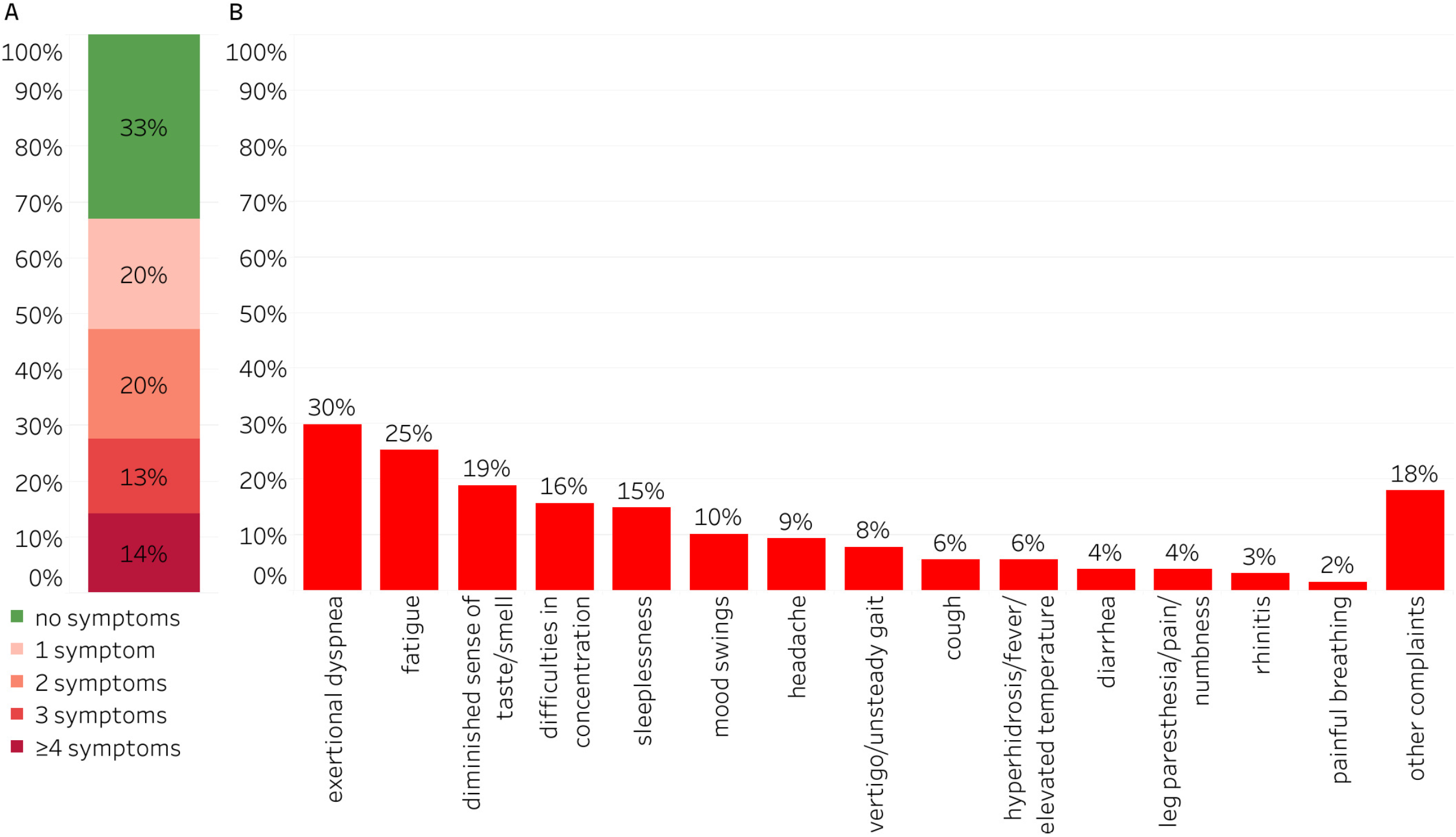
Proportions of participants reporting no, 1, 2, 3, or ≥4 symptoms **(A)** and each individual symptom **(B)** as a consequence of COVID-19, 6 months after the infection

**Figure 2:**
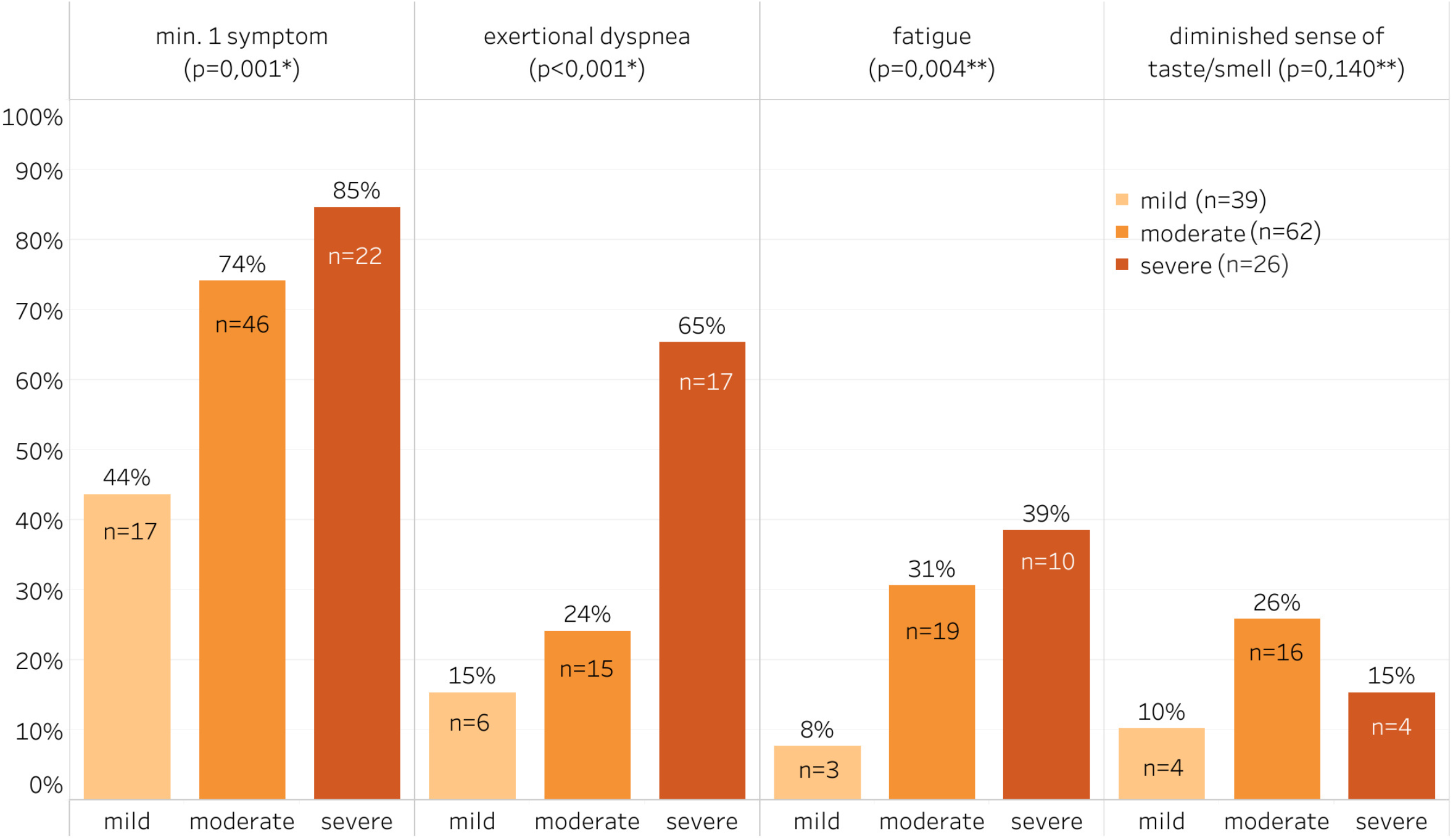
Proportion of participants reporting at least one symptom, exertional dyspnea, fatigue, or diminished sense of taste/smell as a consequence of COVID-19, 6 months after the infection; stratified by severity of the acute illness (mild: asymptomatic or mild symptoms, moderate: strong symptoms with fever or cough but no dyspnea, severe: strong symptoms including dyspnea, oxygen requirement, or mechanical ventilation); Group comparisons with Chi square* / Fisher exact Test**

**Figure 3:**
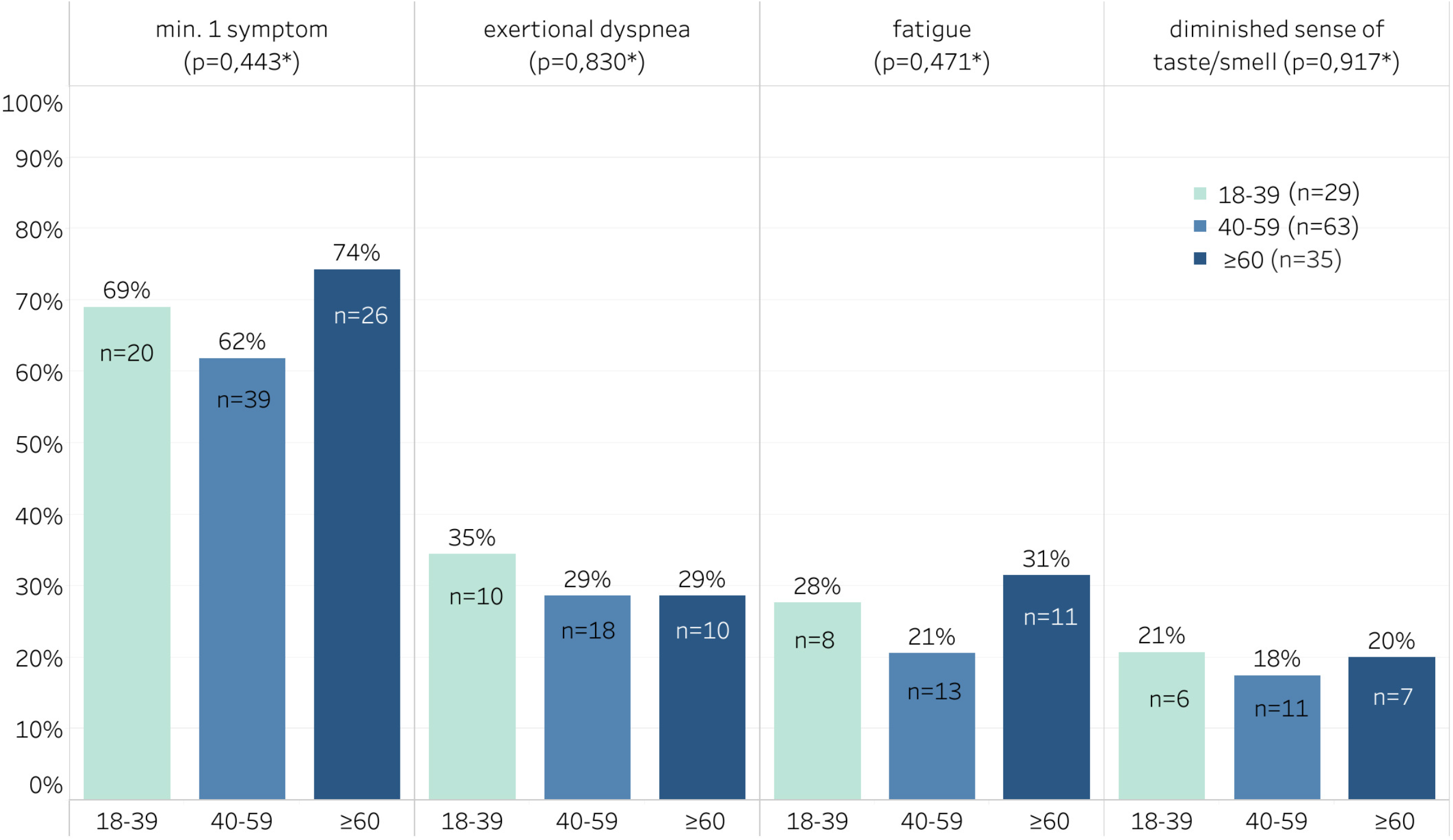
Proportion of participants reporting at least one symptom, exertional dyspnea, fatigue, or diminished sense of taste/smell as a consequence of COVID-19, 6 months after the infection; stratified by age group (18-39 years, 40-59 years, ≥ 60 years); * Group comparisons with Chi square test

**Figure 4:**
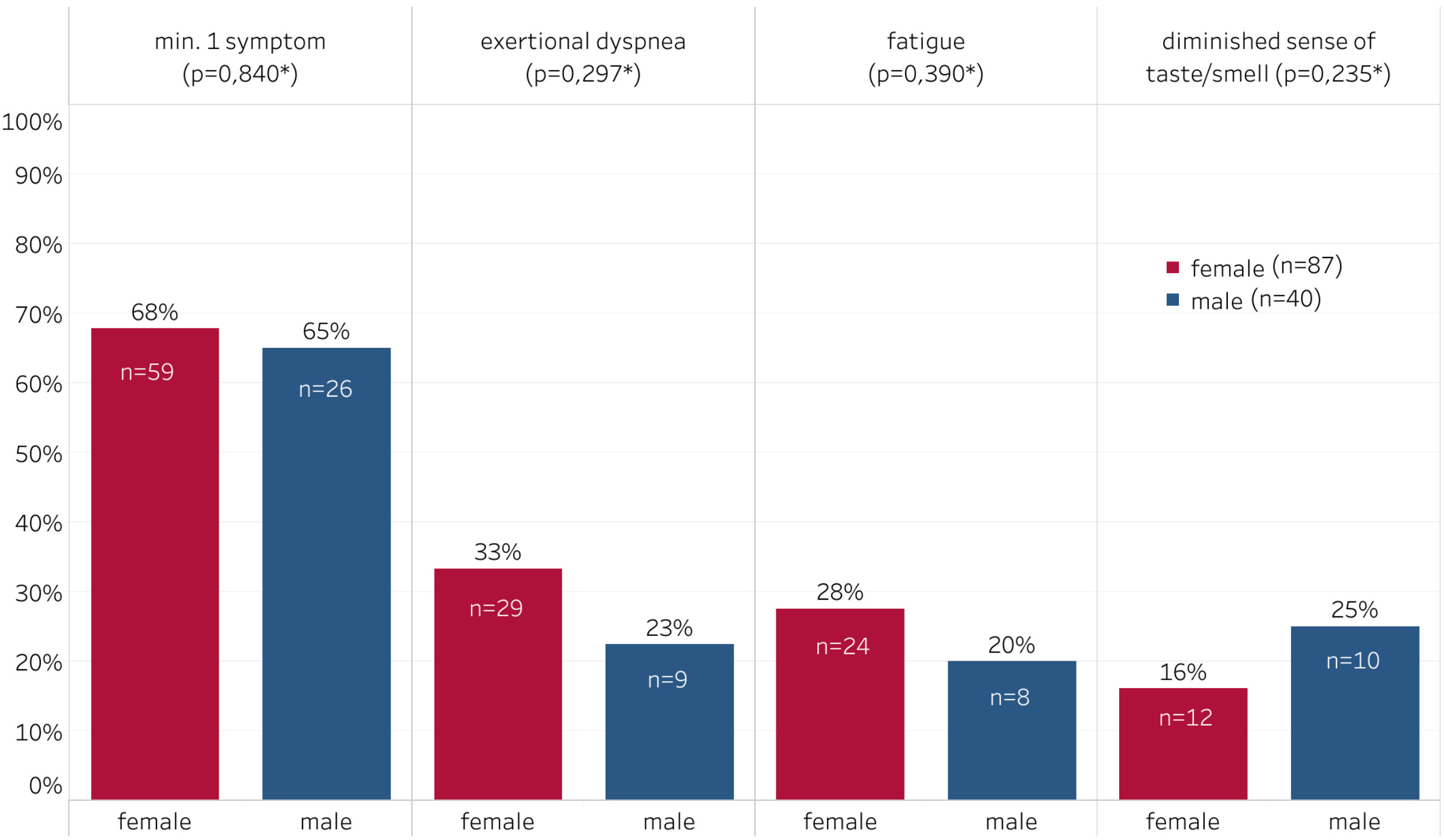
Proportion of participants reporting at least one symptom, exertional dyspnea, fatigue, or diminished sense of taste/smell as a consequence of COVID-19, 6 months after the infection; stratified by sex; * Group comparison with Fisher exact test

## Discussion

We found that symptoms experienced as a consequence of COVID-19 6 months after the infection were common. The most frequent symptoms in our study - exertional dyspnea, fatigue and diminished sense of taste/smell – have been reported previously. However, the pattern of the most common symptoms differed between previous studies and ours (2, 3). For all symptoms analyzed, age group and sex were not associated with symptom-frequency at 6 months.

Persisting exertional dyspnea and fatigue were linked to a more severe course of acute COVID-19 in our study. This link may reflect a higher extent of structural damage during the severe acute illness. Alternatively, it may result from a prolonged period of physical inactivity or necessary medical interventions. Unlike exertional dyspnea and fatigue, diminished sense of taste/smell at 6 months was reported independently from the severity of the acute illness. This symptom may therefore reveal a specific vulnerability of certain individuals.

The observed frequency of symptoms at 6 months is surprisingly high given that we mainly studied individuals treated as outpatients during their acute illness. Therefore, possible causes of a bias for overestimation deserve attention: Firstly, we relied on subjective information provided by the study participants. Therefore, we cannot judge to what extent the reported symptoms would correlate with objective testing of exercise capacity, lung function or smell and taste thresholds. Secondly, participants may have connected unrelated symptoms to COVID-19. Thirdly, recruitment for this online study was not population-based, but through advertisements in news and social media. With an inclusion 0 to 4 months after the acute infection, individuals with ongoing symptoms may have been more likely to participate. However, this study was advertised under its primary aim of examining the associations between lifestyle and the severity of acute COVID-19, not as a study of ‘long COVID’. Additionally, the participants in the prospective follow-up of the study were not more likely to have residual symptoms at baseline than those declining to register for the follow-up (**table 1**). This observation makes the preferential recruitment of individuals with a propensity for long-term symptoms less likely.

### Conclusions

Although we cannot fully exclude overestimation of symptom frequencies in our study, ‘long COVID’ appear be common at 6 months – even after a mild course of the acute illness. To what degree these symptoms result from the viral infection, from therapeutic measures, or from psychosocial factors associated with the acute illness, remains to be determined. Hence, a thorough scientific analysis of the phenomenon of ‘long COVID’ is required. Additionally, complaints of individuals after acute COVID-19 have to be taken seriously by the medical community for better treatment and support.

## Data Availability

Anonymized study data is available from the corresponding author upon reasonable request.

## Acknowledgments

We thank all participants in the *Life&Covid* Study for their important contribution.

## Author Contributions

Conceptualization, all; Formal Analysis, B.R., A.L.; Data Curation, V.S., B.R.; Writing - Original Draft, B.R., A.L.; Writing – Review & Editing, all; Visualization, B.R.; Supervision, A.L.; Project Administration, V.S.; Funding Acquisition A.L., J.S.; A.L. is the guarantor of this work. This work was not published previously.

## Data availability

Anonymized study data is available from the corresponding author upon reasonable request.

